# COVID-19 Growth in Rural versus Urban Counties with Major Universities at the Start of the 2020 Academic Year

**DOI:** 10.1101/2020.11.25.20238642

**Authors:** Mike Penuliar, Candice Clark, Debbie Curti, Cathy Hudson, Billy Philips

## Abstract

Universities play a central role in a rural or small town’s economy. They are often the main forms of enrichment to the lives of the longtime residents, the students, and the employees. Unfortunately, during a global pandemic, the migration and movement of young people in these communities can likely cause a rapid infection spike and drive spread easily, especially relative to larger urban areas. The current study investigates the relationship between COVID-19 case growth, university-county rurality, and time at the beginning of the Fall 2020 academic semester. Findings showed that small metro and non-metro counties with universities had a dramatic infection spike near the beginning of the semester and infection growth remained significantly higher than their large and medium metro counterparts for the duration of the study. Suggestions to slow the spread in rural communities are discussed.

## Introduction

When the novel coronavirus, COVID-19, first appeared in the United States, experts predicted a slow infection rate that would eventually spread from the coastal urban centers to middle and rural America.^1^ Already struggling to overcome their endemic health disparities, rural areas now had to prepare for a pandemic on their horizon with little time, funding, and resources. With poor health literacy and a population consisting of many older Americans with pre-existing conditions, the rural parts of the United States were cited as prime locations for the disease to propagate and thrive.^2^ Adding to the precariousness, many rural counties are also home to major universities, where the mostly transient yet sizeable student demographic interact and share spaces with long-time residents, some of which may be highly vulnerable to COVID-Further, university life often encourages social gatherings and the attending of numerous on- and off-campus events. Taken together, it was only a matter of time before the COVID-19 infection rate soared as universities opened for the Fall 2020 semester.

Despite preventative measures taken by American universities, over 130,000 cases (predominantly students) and more than 70 deaths (primarily employees) have accumulated as of September 2020.^3^ The ravages of COVID-19 in rural university communities are unfortunate as these institutions create jobs and support large portions of their community. And much of the local economy in a rural university town is reliant on influxes of students and university driven tourism. Small and non-metro (SNM) counties often contain a smaller centralized city or town surrounded by high rurality. With limited entertainment, socialization, and errand options, rural residents may opt to go to these central areas for events and amenities. Further, the local workforce outside of the university is populated by many student workers. For these reasons, some may unsuspectingly contract the virus when conducting their affairs about town.

The current study aimed to investigate the relationship between university-county rurality, time, and COVID-19 infection growth at the start of the Fall 2020 academic semester. First, we hypothesized that SNM counties with universities compared to large and medium metro (LMM) counties with universities would have a significant spike in county-wide infection rates at the beginning of the Fall 2020 academic semester, aligning with the rapid migration of students into and within these communities, before settling down a few weeks later. Secondly, because of limited options to conduct one’s business in rural areas and the prospective high amount of county-wide movement and intermingling of the university’s population, we also hypothesized that SNM counties’ infection growth rates would remain higher than LMM counties for the duration of the observation period of the study.

## Methods

All data collected was public. Longitudinal COVID-19 county case data was pulled from *The New York Times’* repository.^6^ Daily percent change in total cases was calculated from August 1, 2020 to September 27, 2020. A 7-day percent change average was calculated for the previous 8 weeks, from September 27, 2020 with Week 1 as August 3, 2020 to August 9, 2020 and Week 8 as September 21, 2020 to September 27, 2020.

Counties were grouped according to the Center for Disease Control – National Center for Health Statistics’ Urban-Rural classification system.^7^ Noncore counties had a population of 9,999 or less. Micropolitan counties had a population between 10,000 and 49,999. Small metro counties had a population between 50,000 and 249,999. Medium metro counties had a population of 250,000 and 999,999. Large metro counties had a population of 1,000,000 or more. Large metro counties were further classified as large fringe metro or large central metro according to the Center for Disease Control (CDC). Large central metro counties are counties that contain the entire population of the large principal city of the metropolitan statistical area (MSA), or are completely contained within the largest principal city of the MSA, or contain at least 250,000 residents of any principal city in the MSA. All others large metro counties that did qualify as a large central metro county were classified as large fringe metro counties.

The Homeland Infrastructure Foundation provided the university enrollment dataset.^8^ Universities with at least 15,000 students enrolled during the 2018-2019 academic year were selected for this analysis. Five majority online universities were removed after segmenting. These removed universities had an enrollment range of 79,152 to 121,437. Counties were then classified on the criteria of the presence of at least one university with 15,000+ students enrolled and were then analyzed using a repeated measures design.

## Results

Of the counties containing universities with 15,000+ students enrolled, there were a total of 49 large central metro counties, 31 large fringe metro counties, 49 medium metro counties, 29 small metro counties, 14 micropolitan counties, and 1 non-core county. Due to the presence of only a single non-core county and to enhance further analyses, micropolitan and non-core counties were combined to form the super-ordinate non-metro county category as described by the CDC.

An 8 (week) x 5 (county type) repeated measures ANOVA was ran. See Table 1 for means and standard deviations. The analysis found a significant within-subjects main effect for week, *F*(3.24,544.88) = 16.92, *p* < .001, *η*_*p*_^2^ = .091. There was also a significant between-subjects main effect for county type, *F*(4, 168) = 26.07, *p* < .001, *η*_*p*_^2^ = .383. Importantly, a significant week x county type interaction qualified all lower order main effects *F*(12.97,544.88) = 8.87, *p* < .001, *η*_*p*_^2^ = .174.

**Table 1.**
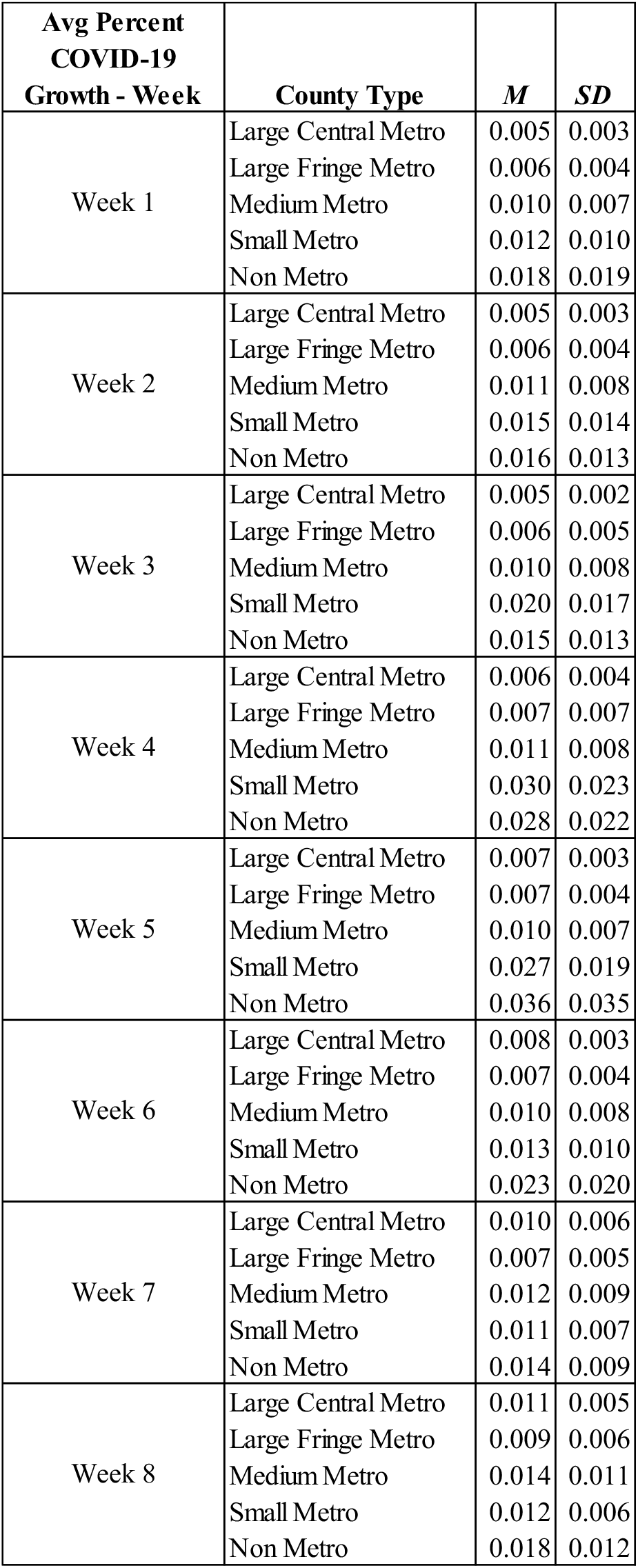
Means and Standard Deviations Across Week and County Type

| **Avg Percent**<br>**COVID-19**<br>**Growth - Week** | **County Type** | ***M*** | ***SD*** |
| --- | --- | --- | --- |
| Week 1 | Large Central Metro | 0.005 | 0.003 |
|  | Large Fringe Metro | 0.006 | 0.004 |
|  | Medium Metro | 0.010 | 0.007 |
|  | Small Metro | 0.012 | 0.010 |
|  | Non Metro | 0.018 | 0.019 |
| Week 2 | Large Central Metro | 0.005 | 0.003 |
|  | Large Fringe Metro | 0.006 | 0.004 |
|  | Medium Metro | 0.011 | 0.008 |
|  | Small Metro | 0.015 | 0.014 |
|  | Non Metro | 0.016 | 0.013 |
| Week 3 | Large Central Metro | 0.005 | 0.002 |
|  | Large Fringe Metro | 0.006 | 0.005 |
|  | Medium Metro | 0.010 | 0.008 |
|  | Small Metro | 0.020 | 0.017 |
|  | Non Metro | 0.015 | 0.013 |
| Week 4 | Large Central Metro | 0.006 | 0.004 |
|  | Large Fringe Metro | 0.007 | 0.007 |
|  | Medium Metro | 0.011 | 0.008 |
|  | Small Metro | 0.030 | 0.023 |
|  | Non Metro | 0.028 | 0.022 |
| Week 5 | Large Central Metro | 0.007 | 0.003 |
|  | Large Fringe Metro | 0.007 | 0.004 |
|  | Medium Metro | 0.010 | 0.007 |
|  | Small Metro | 0.027 | 0.019 |
|  | Non Metro | 0.036 | 0.035 |
| Week 6 | Large Central Metro | 0.008 | 0.003 |
|  | Large Fringe Metro | 0.007 | 0.004 |
|  | Medium Metro | 0.010 | 0.008 |
|  | Small Metro | 0.013 | 0.010 |
|  | Non Metro | 0.023 | 0.020 |
| Week 7 | Large Central Metro | 0.010 | 0.006 |
|  | Large Fringe Metro | 0.007 | 0.005 |
|  | Medium Metro | 0.012 | 0.009 |
|  | Small Metro | 0.011 | 0.007 |
|  | Non Metro | 0.014 | 0.009 |
| Week 8 | Large Central Metro | 0.011 | 0.005 |
|  | Large Fringe Metro | 0.009 | 0.006 |
|  | Medium Metro | 0.014 | 0.011 |
|  | Small Metro | 0.012 | 0.006 |
|  | Non Metro | 0.018 | 0.012 |

According to Figure 1, non-metro and small metro counties trended similarly with large increases in their COVID-19 case growth from Weeks 2 to 6. This finding satisfied our hypothesis as this was when student populations were moving back to their university communities. All other groups remained mostly flat in their weekly average growth. Interestingly, from Week 6 to 8, non-metro counties increased in their growth rate, but small metro counties continued to decrease. Marginal means analyses were then performed (Table 2). Aligning with our second hypothesis, starting at week 3 to the end of the study period, SNM compared to LMM saw significantly large case growth week over week, never returning to the growth rate prior to the student influx.

**Table 2.** Medium Metro and Larger versus Small Metro and Smaller – Means and Standard Deviations

| **Week** | **County Type** | ***M*** | ***SD*** | **99%**<br>**CI LB** | **99%**<br>**CI UB** |
| --- | --- | --- | --- | --- | --- |
| 1 | Medium Metro and Larger | 0.012 | 0.011 | -0.006 | 0.002 |
|  | Small Metro and Smaller | 0.014 | 0.007 |  |  |
| 2 | Medium Metro and Larger | 0.010 | 0.011 | -0.005 | 0.001 |
|  | Small Metro and Smaller | 0.012 | 0.007 |  |  |
| 3* | Medium Metro and Larger | 0.008 | 0.011 | -0.012 | -0.004 |
|  | Small Metro and Smaller | 0.016 | 0.007 |  |  |
| 4* | Medium Metro and Larger | 0.008 | 0.011 | -0.028 | -0.016 |
|  | Small Metro and Smaller | 0.030 | 0.013 |  |  |
| 5* | Medium Metro and Larger | 0.008 | 0.011 | -0.027 | -0.016 |
|  | Small Metro and Smaller | 0.030 | 0.013 |  |  |
| 6* | Medium Metro and Larger | 0.007 | 0.011 | -0.015 | -0.007 |
|  | Small Metro and Smaller | 0.018 | 0.007 |  |  |
| 7* | Medium Metro and Larger | 0.008 | 0.011 | -0.012 | -0.004 |
|  | Small Metro and Smaller | 0.016 | 0.007 |  |  |
| 8* | Medium Metro and Larger | 0.007 | 0.011 | -0.011 | -0.003 |
|  | Small Metro and Smaller | 0.014 | 0.007 |  |  |

\* = The mean difference is significant at $p < .01$ level, Bonferonni adjusted

**Figure 1.**
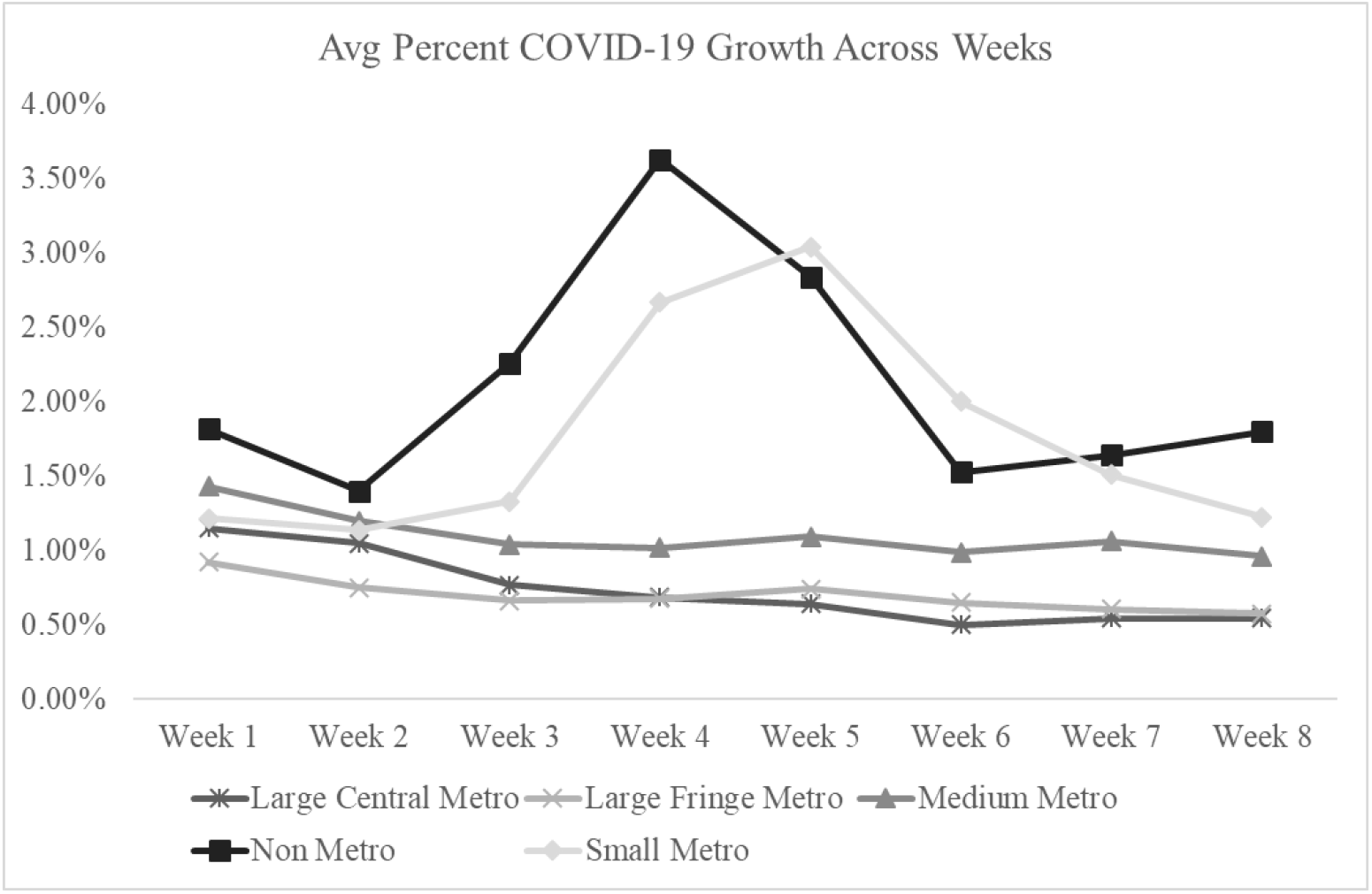
Average Covid-19 Growth Rate Across Week and County Type

## Discussion

It is clear that there is a relationship between university rurality and COVID-19 growth. SNM counties with major universities experienced an intense spike in COVID-19 case growth between Week 3 and 6 in the study: August 17, 2020 to September 13, 2020. Further, when comparing SNM counties against LMM counties, the former grew at a significantly faster pace from Week 3 to the end of the observation range, September 27, 2020. These findings align with our prediction that once university students started moving back into these communities and congregating throughout the county, they likely helped spur the spread of COVID-19 within their university population and the long-time and potentially vulnerable residents of the county.

Related research found that COVID-19 spring 2020 infection rates were substantially higher among young and middle-aged adults in rural compared to urban areas,^9^ which may be attributed to universities as detailed in this study. The CDC also reported that from August 2020 to September 2020 cases among those aged 18-22 increased by 55% nationally.^10^ Though early fall 2020 COVID-19 growth has mostly been attributed to young people, mortality rate remains highest in older adults and those with preexisting conditions.^11^ And high overall caseloads in rural counties have the potential to make their way to those more vulnerable and drive unneeded suffering.

These results are vital because as rurality increases, resources decrease and population vulnerabilities increase.^12,13^ Specifically, rural counties have fewer and older physicians, older patients with less income and insurance, and poorer internet infrastructure for telemedicine. These factors have the potential to worsen pandemic responses and patient prognoses. Many rural healthcare systems are also considered under resourced and may be situated a significant commute time away from residents. If university populations truly play a role in COVID-19’s growth in SNM counties, there is much to be done to help protect the rural vulnerable.

Emphases on face coverings, surveillance testing, contact tracing, and online schooling options are necessary lines of defense. Unfortunately, a recent analysis found that more than 66% of colleges do not have an adequate testing methodology in place, and many of these universities are also located in counties with escalating COVID-19 growth as of this writing.^14^ It is key to educate the community on the social determinants of health and depoliticize good public health behaviors in order to keep this disease from further decimating rural communities. Finally, improving telemedicine infrastructure will be integral for many during this pandemic. Many communities have responded and performed admirably. Though progress has been made, vulnerable populations are still in danger as cases and deaths continue to climb in rural university counties and rural areas overall. There is still much work to be done so those in under resourced rural communities can have an equitable chance of fighting this virus.

## Data Availability

All data collected was public. Longitudinal COVID-19 county case data was pulled from The New York Times' repository. Counties were grouped according to the Center for Disease Control - National Center for Health Statistics' Urban-Rural classification system. The Homeland Infrastructure Foundation provided the university enrollment dataset

https://github.com/nytimes/covid-19-data

https://www.cdc.gov/nchs/data_access/urban_rural.htm

https://hifld-geoplatform.opendata.arcgis.com/datasets/colleges-and-universities-campuses

